# SARS-CoV-2 infection in Ivory Coast: a serosurveillance survey among gold mine workers

**DOI:** 10.1101/2021.01.27.21249186

**Authors:** Jean Marie Milleliri, Daouda Coulibaly, Blaise Nyobe, Jean-Loup Rey, Franck Lamontagne, Laurent Hocqueloux, Susanna Giaché, Antoine Valery, Thierry Prazuck

## Abstract

**Background:** Eight months after the detection of the first COVID-19 case in Africa, 1,262,476 cases have been reported in African countries compared to 72 million worldwide. The real burden of SARS-CoV-2 infection in West Africa is not clearly defined. The aim of the study was to evaluate the seroprevalence of SARS-CoV-2 in half of the 3,380 workers of several mining companies operating in two mines in the Ivory Coast and having its headquarters in the economic capital Abidjan.

**Methods:** From 15th July to 13th October 2020, a voluntary serological test campaign was performed in the 3 sites where the companies operate: two mines, and the headquarters in Abidjan.We performed a COVID-PRESTO rapid test for the detection of IgG and IgM on capillary blood. A multivariate analysis was performed to identify independent sociodemographic characteristics associated with a higher SARS-CoV-2 seroprevalence rate.

**Results:** A total of 1,687 subjects were tested. 91% were male (n= 1,536) and mean age was 37 years old. The overall crude seroprevalence rate was 25.1% (n=422), but differing significantly between different sites, rising from 13.6% (11.2%-16.1%) in mine A to 34.4% (31.1%-37.7%) in mine B and 34.7% (26.2%-43.2%) in Abidjan. Non-resident workers in mines had a significantly lower prevalence rate than those living full-time in mines. Seroprevalence was 26.5% in natives of the Ivory Coast, while people coming from countries other than Africa were less likely to be SARS-CoV-2 seropositive. Among the 422 positive subjects, 74 reported mild symptoms in the three previous months and one was hospitalized for a severe COVID-19 infection.

**Conclusion:** The prevalence of SARS-CoV-2 infection among mine workers in Ivory Coast is high. The low morbidity observed has probably led to an underestimation of the burden of this infection in West Africa. The high prevalence reported in subjects living in Abidjan, who have not any close contact with mine workers, may be indicative of the real seroprevalence in the Ivory Coast capital.

---

CoronaVirus Disease 2019 (COVID-19) represents an unprecedented international public health challenge responsible for significant morbidity and mortality. In March 2020, the World Health Organization (WHO) officially declared the COVID-19 pandemic. As of 17th December 2020, the Severe Acute Respiratory Syndrome CoronaVirus 2 (SARS-CoV-2) caused 1,637,000 deaths out of more than 72 million cases reported worldwide. Unfortunately these numbers continue to increase every day^1^.

Officially, according to the latest October 21^st^ 2020 WHO report, COVID-19 cases in Africa were 1,262,476 and COVID-19 related deaths were 28,601, with no significant change in incidence over weeks (29,919 new confirmed cases and 474 new deaths from 46 countries, compared to 30,145 cases and 747 deaths in the previous week)^2^.

Considering the daily increase in the number of COVID-19 cases all around the world, it appears that Africa is controlling the epidemic spread better than Europe, United States or South America. Catastrophic scenarios about potentially devastating effect of COVID-19 in Africa have fortunately not yet occurred. The reasons are multiple and not always clearly established^3^. Some authors argue that this epidemic reached Africa a few weeks after Europe and African leaders adopted preventive measures thanks to acquired knowledge of what was happening in Europe, having probably learned important lessons from previous major outbreaks, like 2014 Ebola virus disease in West Africa. In addition, many parts of Africa are still isolated and not linked to significant tourism and international business related traffic. However, according to recent data, SARS-CoV-2 serological positivity in South Africa appears to be so high that it approaches the herd immunity threshold (i.e. more than 60%) (Jonny Myers, Maverick Citizen, Open Edition, 7 oct 2020)^4^. Did SARS-CoV2 infection largely and silently spread in Africa, or did factors such as environment, younger population, climate and policies limit the penetration of the virus in most African countries?

Since March 2020, Ivory Coast, Mali and Burkina Faso have faced the SARS-CoV-2 outbreak almost simultaneously. In this report, we present the results of a COVID-19 serological testing campaign conducted in Ivory Coast among employees of a gold mining company using a reliable rapid Point-of-Care (POC) antibody test.

## Background

Several mining companies operate in West Africa. Occupational Health department is in charge of mine workers as well as administrative staff. Mines are mainly located in rural areas of the country, between 200 km and 500 km far from the economic capital of the country (Abidjan) where these companies’ headquarters are located. Three-thousand three hundred and eighty people are currently working in these sites, including the Abidjan headquarters. Of these 3,380 workers, 1,360 are direct employees and 2,020 are subcontractor’s employees of different companies. Direct employees working in the mines live on site in a rather confined place, while the subcontractor’s employees come back home in their villages/towns every day. Administrative staff and other workers located in Abidjan headquarters are mainly exposed to COVID-19 in their daily activities in contact with the Ivorian urban population of Abidjan. The objective of this testing campaign was to sensitize and protect employees and their families, particularly when a recent SARS-CoV-2 infection was diagnosed.

## Subjects and methods

The serological screening based on voluntary testing was conducted in the period between July 15^th^ and October 13^th^. All employees were informed about the objectives of the campaign and were invited to participate. No clinical criteria were required to be tested. Tests were performed at the different work sites by healthcare personnel according to the manufacturers’ instructions after a two-hours training session. For each participant, a written informed consent was obtained. Test result was provided within 15 minutes. A photograph of each cassette was systematically performed to be assessed by an independent operator. A standardized questionnaire was filled for each subject, including age, sex, working status and previous symptoms potentially related to a SARS-CoV-2 infection in the previous five months (i.e. fever, dyspnea, cough, flu-like syndrome, anosmia/ageusia).

### Point-of-care used test

The COVID-PRESTO® SARS-CoV-2 IgG/IgM antibody test kit is a lateral flow immune-chromatographic assay targeting antibodies specific to the SARS-CoV-2 N-protein. It has been manufactured and marketed by AAZ-LMB and already approved by French authorities. This rapid diagnostic test (RDT) uses anti-human IgM antibodies (test line IgM), anti-human IgG antibodies (test line IgG) and rabbit IgG (control line C) immobilized on a nitrocellulose strip. The Conjugate (recombinant SARS-CoV-2 antigens labeled with colloidal gold) is also integrated into the strip. When a specimen is well placed in the sample and the assay buffer is added, the IgM and IgG antibodies, if present, bind to the SARS-CoV-2 conjugates forming an antigen-antibodies complex. This complex migrates through a nitrocellulose membrane by capillary action. When the complex meets the line of the corresponding immobilized antibodies (anti-human IgM and/or anti-human IgG), the complex is trapped and form a burgundy-colored band confirming test result. Results must be read within 10 minutes by two independent operators. When the control line is the only one present, the sample is negative. If the control line does not appear, the test is invalid and should be repeated with a new cassette. Performances of this test have already been assessed in real life, regardless of the manufacturer’s instructions. The sensitivity of COVID-PRESTO® test ranged from 10% for patients having experienced their first symptoms from 0 to 5 days earlier to 100% in patients whose symptoms occurred more than 15 days earlier^5^.

### Statistical analysis

Data were collected on an Excel spreadsheet. Data were categorized by mine site, sex, age and working characteristics (i.e. direct employees vs subcontractor’s employees). Qualitative and quantitative variables including the proportion of seropositivity were reported in percentage and a 95% confidence interval. Multivariate logistic regression was used to determine the relationship between demographics characteristics and seropositivity to the SARS-CoV-2. A p value of less than 0.05 was regarded as statistically significant. Only statistically significant variables were introduced in the multivariate model. Analysis was carried out using R studio software.

### Ethical considerations

The GISPE Research and Ethical committee approved the survey on May 15^th^ 2020 (N° EC-GISPE014). Potential participants were informed about the campaign purpose and procedures.

## Results

Among the 3,380 employees, a total of 1,687 subjects (49.9%) were included in the study. One thousand five hundreds thirty-five were male and 151 were female. Mean age was 37 (18-66) years old; 1,566 employees (92.7%) worked in the mine sites and 121 (7.3%) in Abidjan. Among mining workers, 845 were direct employees living night and day in the compound (587 in mine A and 258 in mine B,), and 718 were subcontractor’s employees of other companies working in the mine during the day but coming back to village after work (213 in mine A and 501 in mine B). One thousand four hundreds fifty-eight workers were native from Ivory Coast (86.4%), 143 from another African country (8.5%) and the remaining 87 (5.1%) came from 12 worldwide countries (Details in Table I) Subjects working in Abidjan were significantly older (63% over 36 years old) and the proportion of male workers was half compared to mine sites. Among the 1,687 people included in the study, 422 had a positive serological test (either IgG and/or IgM). Overall seroprevalence was 25.1% and prevalence rates were similar when stratified by age or gender. According to the different working sites, the seroprevalence was 34.4%, 34.7%, and 13.6% in mine A, Abidjan headquarters and mine B. Seroprevalence rate was significantly lower in subcontractors workers than in those living full time in the mines. Ivory Coast natives had the highest prevalence rate [26.5% (24.3% - 28.8%)]. Conversely, people coming from countries outside Africa were less likely to be SARS-CoV-2 seropositive (Details in Table 2).

**Table I:**
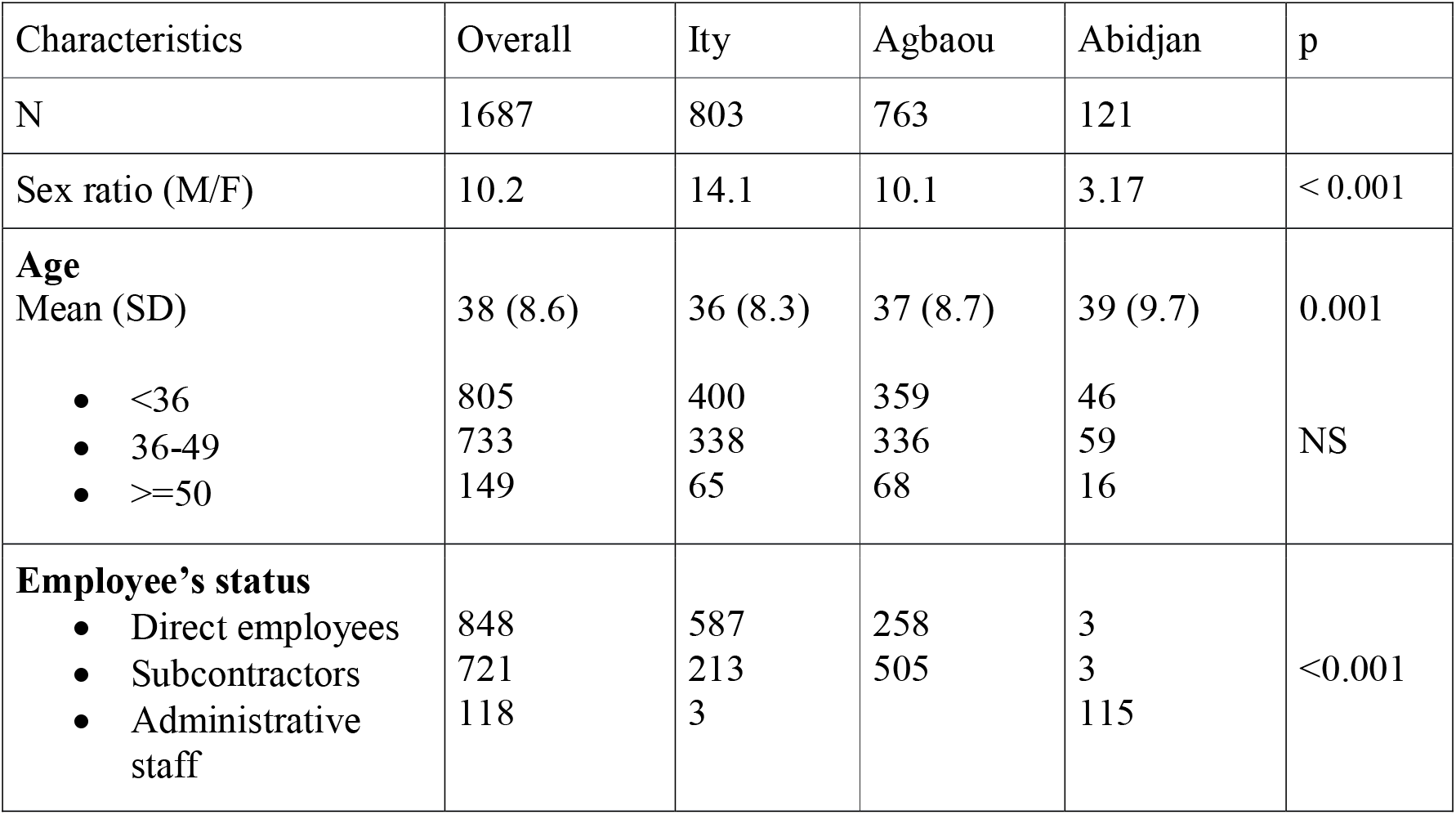
Sociodemographic characteristics of the study population

| Characteristics | Overall | Ity | Agbaou | Abidjan | p |
| --- | --- | --- | --- | --- | --- |
| N | 1687 | 803 | 763 | 121 |  |
| Sex ratio (M/F) | 10.2 | 14.1 | 10.1 | 3.17 | < 0.001 |
| **Age**<br>Mean (SD) | 38 (8.6) | 36 (8.3) | 37 (8.7) | 39 (9.7) | 0.001 |
| • <36 | 805 | 400 | 359 | 46 | NS |
| • 36-49 | 733 | 338 | 336 | 59 |  |
| • >=50 | 149 | 65 | 68 | 16 |  |
| **Employee's status** |  |  |  |  | <0.001 |
| • Direct employees | 848 | 587 | 258 | 3 |  |
| • Subcontractors | 721 | 213 | 505 | 3 |  |
| • Administrative staff | 118 | 3 |  | 115 |  |

**Table II:**
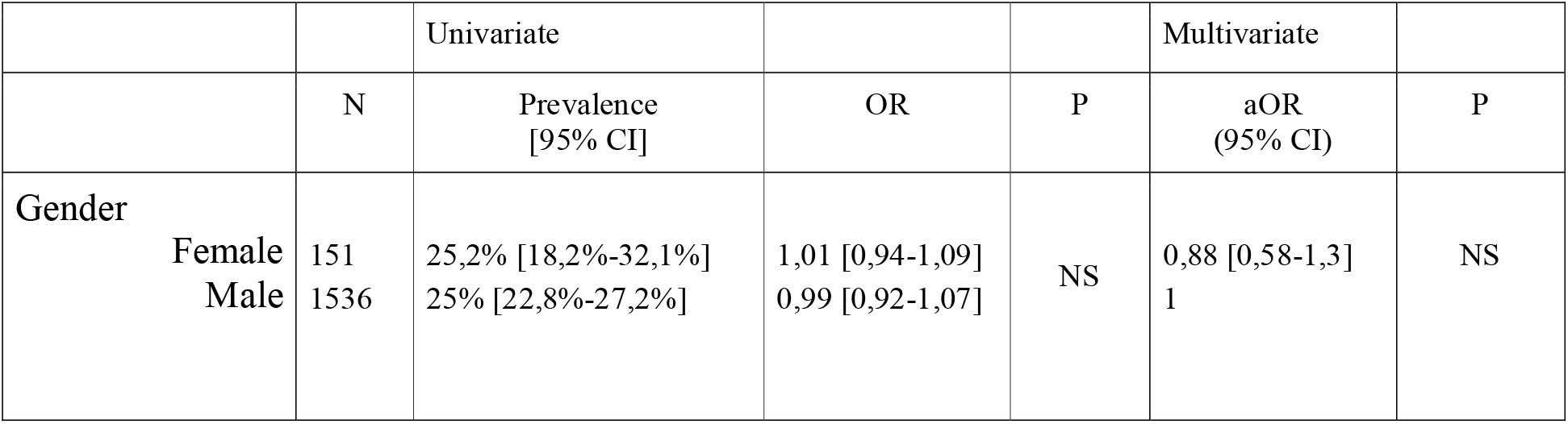
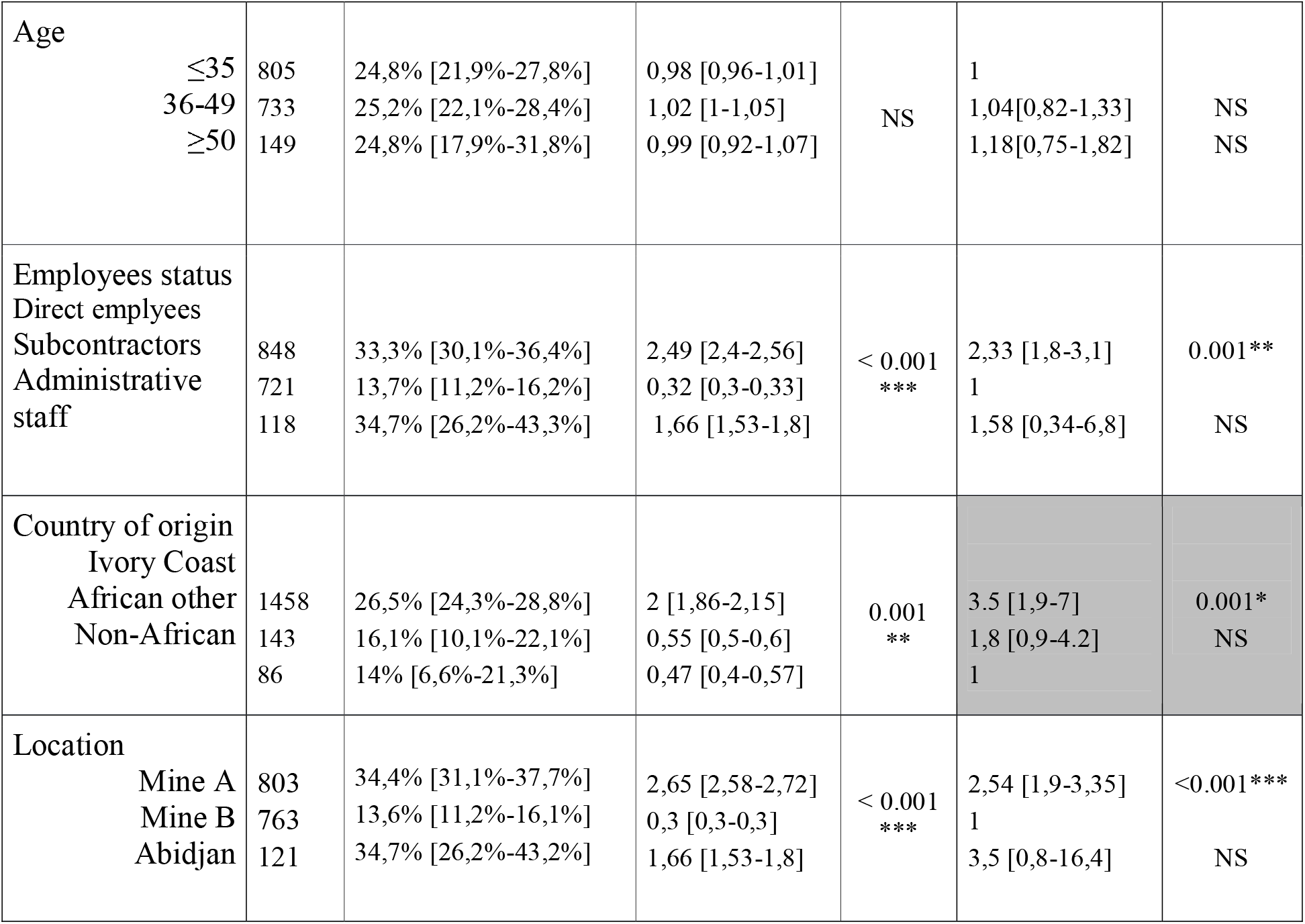
SarS-CoV2 seroprevalence rates, odds ratios and 95% confidence interval according to sociodemographic characteristics plus adjusted odds ratio (logistic model).

Among the 422 seropositive subjects, only one had previously been hospitalized for a suspected COVID-19. Eighteen additional subjects had mild symptoms and were diagnosed by a positive real-time reverse transcription polymerase chain reaction (RT-PCR) test. Among the 404 remaining subjects, 74 reported mild nonspecific symptoms that may or may not be attributable to a SARS-CoV-2 infection, particularly in a tropical setting, but were not tested at the time of symptoms.

## Discussion

The current epidemiological study conducted on a specific African population reports a SARS-CoV-2 seroprevalence as high as 35% in gold mining workers as well as in administrative staff living in Abidjan. This high seroprevalence rate has been reached before July 2020, and it remained stable after the first pandemic wave, between July and October 2020. This silent outbreak had not previously been detected because most cases were asymptomatic.

From March to December 1^st^, 2020, Ivory Coast reported 21,250 SARS-CoV-2 infections and 127 COVID-19 related deaths, meaning that only the 0.084% of Ivory Coast’s 25 million inhabitants has been infected ^2^. Official available data do not reflect the burden of infection but may reflect the emerging part of the iceberg. To our knowledge, there are very few published studies evaluating SARS-CoV-2 seroprevalence in African countries. In Togo, among 955 participants from five different sectors, the prevalence rate was 0.9% (95% CI: 0.4%-1.8%)^6^. In a study conducted in Kenya from April to June 2020 on 3,174 blood donors’ samples, seroprevalence was 5.2% (95%CI: 3.7%-7.1%)^7^. In two small populations of household contacts and healthcare workers in Nigeria, seroprevalence rates rise to 25.4% and 45% respectively ^8,9^. Among the available pre-print or published studies, seroprevalence was evaluated using Rapid Diagnostic Tests (RDTs) in three studies, enzyme-linked immunosorbent assay (ELISA) in three studies and Clinical Laboratory Improvement Amendments (CLIA) in one study^6–12^. A recent study aiming to detect the persistence of SARS-CoV-2 antibodies 3 to 4 months after the onset of symptoms in healthcare workers at the Strasbourg University hospital reported that ELISA serological test has a 59% sensitivity and Biosynex RDT a 85% sensitivity compared to the S-flow reference test developed by the French National Reference Center for SARS-CoV-2 infection (Pasteur Institute, Paris, France)^13^. The RDT used in our study showed high sensitivity even more than 21 days after onset of symptoms, as well as a 100% specificity in a study carried out independently from the manufacturer^14^. Hence, seroprevalence rates from studies using ELISA tests must be cautiously interpreted as they may underestimate the real data.

In our study, 14% to 35% of workers were SARS-CoV-2 seropositive, despite having no symptoms or at least no severe disease. These data appear to be quite different than that observed in the rest of the world, but similar results have already been described in Ibadan, Nigeria, in healthcare workers who are much more exposed to the infection than mine workers ^9^. A high level of seropositivity was found in both mining workers and administrative staff living in the economic capital. In mine A, as people live together, a cluster phenomenon may have occurred, explaining the difference in seroprevalence with mine B, which was more confined during the first wave with limited circulation between mine B and Abidjan. Subjects living in mine B had the same life habits as the African population living in Abidjan. Therefore, we can expect that this similar seroprevalence reflects the real one in Abidjan.

In mine B, subcontractors’ workers constitute 67% of the working population compared to 29% in mine A. As subcontractors’ worker do not live full-time in the mine, the lower seroprevalence of this population, could reflect a lower prevalence in rural areas of the country.

In the capital, seropositivity was 34.7% among the 121 subjects working in the administrative staff in Abidjan and having no contact with gold mines. This data could reflect the high SARS-CoV-2 seroprevalence in the general population living in Abidjan.

Nevertheless, the most surprising results of the current study are the extremely low percentage of symptomatic cases and mortality rate. Some authors speculate that African SARS-CoV-2 seroprevalence data could be related to a younger population, a favorable climate and possible preexisting immunity due to previous exposure to other coronaviruses ^14^. In our study, mean age was 37 years old and there was no increase in symptoms incidence or hospitalization according to age range. Our prevalence rates do not support hypothesis of a previous cross-reacting immunity due to other coronaviruses.

Relatively stable prevalence rates observed between July and September may indicate that SARS-CoV-2 outbreak occurred as early as March or April as in Europe, mainly not to say exclusively from imported cases, and spread silently but rapidly across the country. The plateau observed since July could be related either to complete travel restrictions, to the end of the first epidemic wave as in Europe, to the nationwide control of the SARS-CoV-2 circulation, or may reflect other phenomena including pre-existing immunity, genetic factors … in the remaining population.

This study has some limitations. Firstly, the included population is not representative of the general Ivoirian population, although subjects living in Abidjan have similar habits to those of the Abidjan’s population. Furthermore, the voluntary test may have selected people thinking to be more exposed to the virus, for example people who previously had mild symptoms or had households contacts of a confirmed SARS-CoV-2 case. Third, even if COVID-PRESTO® has an excellent diagnostic performance, diagnostic sensitivity decreases a few months after the infection, compared to the reference test S-Flow, and may therefore have underestimated the real seroprevalence.

In conclusion, the high SARS-CoV-2 prevalence rate among mining workers recruited for this study confirms a higher proportion of asymptomatic cases than that observed in Europe, South America or United States of America till now. Herd immunity could be achieved in Africa much easier than expected without generating a dramatic health crisis.

## Data Availability

Excel file containing data is available on demand

